# Factors driving switching from vaping of e-cigarettes to smoking tobacco and initiation of multi-inhalant use

**DOI:** 10.1101/2025.03.04.25323341

**Authors:** Enny Banjo, Zoya Ahadian, Nikita Kasaraneni, Howard Chang, Sarala Perera, Kristen Emory, Laura E. Crotty Alexander

## Abstract

The prevalence of e-cigarette use in adolescents is concerning, particularly because of high rates of vapers transitioning to combustible tobacco. It is unclear why young, never smoker, e-cigarette vapers decide to start smoking cigarettes, but the transition may be influenced by perceptions of health risks and accessibility. We designed a social media questionnaire to assess factors driving switches from vaping to smoking, smoking to vaping, and multi-inhalant use. Among respondents (n=864) who switched from vaping to smoking (n=104), primary reasons included: 1. perception that smoking was healthier (44%); and 2. greater accessibility of cigarettes (40%). For switchers from smoking to vaping (n=178), predominant motivations included: 1. having friends or family who vape (40%); and 2. perception that vaping was healthier (36%). Among multi-inhalant users (n=223), key factors driving this behavior included: 1. increased enjoyment (47%); and 2. greater variety (42%). Our findings imply that there is no single, dominant reason driving switching from vaping to smoking. Further, tobacco users are receiving mixed messaging, leading many to believe that combustible tobacco is the healthier option. Public health interventions are needed to prevent initiation of e-cigarette vaping by youth and education of the public about health effects across tobacco products.

## 1. Introduction

When e-cigarettes were introduced to the United States in 2007, they were marketed as a safe alternative to combustible cigarettes. Tobacco use in adolescents had been constantly decreasing, but with the introduction to the market of the fruity, sweet and minty flavored e-cigarettes, tobacco use rapidly up-ticked[1], with use by middle school (ages 10-14), high school (ages 14-18), and college students (ages 19-23) surpassing that among adults[2]. For example, 18.6% of college students between the ages of 19 and 22 have reported using e-cigarette within the past 30 days[3]. Amidst this surge, numerous studies have deliberated on whether e-cigarette use among adolescents poses a risk factor for subsequent cigarette smoking—a phenomenon often termed the gateway effect[4]. According to this theory, e-cigarettes may function as a nicotine starter, leading to nicotine dependence and subsequent use of other tobacco products[4]. Multiple meta-analyses have confirmed that initiation of nicotine e-cigarettes by adolescents increases subsequent use of combustible tobacco[1]. Data thus far also suggests that initiation of e-cigarettes leads to multi-inhalant use of vapes with combustible tobacco and cannabis[5], a pattern of inhalant use that has more adverse effects than either inhalant alone[3, 4]. Most notable, it remains unknown as to why never smokers who vape e-cigarettes begin smoking conventional tobacco.

In this study, we assess the driving forces underlying initiation of combustible tobacco by e-cigarette vapers. We posit that nicotine addiction plays a central role in switching from vapes to cigarettes and transitions to multi-inhalant use. We hypothesize that other factors also contribute, such as lower cost and higher accessibility of cigarettes. Furthermore, we explore other patterns of inhalant usage, including transitions from cigarette smoking to vaping nicotine, multi-inhalant use, and cessation of both habits. We anticipate that individuals may opt for vaping over smoking due to perceived health benefits, while those engaging in multi-inhalant use may seek heightened nicotine intake. Finally, we hypothesize that the primary driver for quitting both e-cigarettes and combustible cigarettes is health benefits.

By examining inhalant use patterns and motivations underlying the choices tobacco users are making, we hope to obtain a deeper understanding of the complex dynamics surrounding e-cigarette and cigarette use, shedding light on potential avenues for interventions, public health outreach, and harm reduction strategies.

## 2. Materials and Methods

### Survey Design and Distribution

A 33-item questionnaire was designed in Survey Monkey, with inhalant questions based on our published University of California San Diego (UCSD) Inhalant Questionnaire[5-7], and the University of Colorado Anschutz 2021 Healthy Kids Survey[8]. Novel questions probing behavioral drivers underlying switching and adding in of combustible tobacco in vapers were also included. UCSD institutional review board (IRB 160204) approval was obtained, and all survey takers gave consent (>18 years of age) or assent (<18 years of age).

The focused survey was posted on multiple social media platforms, including Facebook, Craigslist, and Instagram. To avoid and reduce bot responses, Twitter and Reddit were not used. Individuals completed two bot-detection questions within the survey. A total of 1271 subjects, composed of nonsmokers, e-cigarette users, smokers, and multi-inhalant users, completed our nationwide online survey, with 864 non-bot survey responses identified (Table 1). All survey takers were entered into a random drawing for a $100 gift card.

**Table 1.** Demographic data for survey responders.

|  | All<br>(609)<br>N (%) | Switched:<br>Vaping to<br>Smoking<br>(104)<br>N (%) | Switched:<br>Smoking to<br>Vaping<br>(178)<br>N (%) | Dual Use<br>(223)<br>N (%) | Quit<br>Tobacco<br>(93)<br>N (%) | Other<br>(11)<br>N (%) |
| --- | --- | --- | --- | --- | --- | --- |
| **Age** |  |  |  |  |  |  |
| 16 – 21 years | 11.8 (72) | 14.4 (15) | 9.6 (17) | 11.2 (25) | 15.1(14) | 9.1 (1) |
| >21 years | 88.2 (537) | 85.6 (89) | 90.4 (161) | 88.8 (198) | 84.9 (79) | 90.9 (10) |
| **Gender** |  |  |  |  |  |  |
| Female | 30.0 (183) | 35.6 (37) | 28.1 (50) | 29.1 (65) | 30.1 (28) | 27.3 (3) |
| Male | 58.8 (358) | 51 (53) | 60.7 (108) | 58.3 (130) | 64.5 (60) | 63.6 (7) |
| Transgender Female | 3.8 (23) | 4.8 (5) | 3.9 (7) | 4.0 (9) | 2.2 (2) | 0 (0) |
| Transgender Male | 6.7 (41) | 8.7 (9) | 5.6 (10) | 8.5 (19) | 3.2 (3) | 0 (0) |
| Gender queer/non-binary | 0.7 (4) | 0 (0) | 1.7 (3) | 0 (0) | 0 (0) | 9.1 (1) |
| **Race/Ethnicity\*** |  |  |  |  |  |  |
| Hispanic or Latino | 8.0 (49) | 2.9 (3) | 11.2 (20) | 7.2 (16) | 9.7 (9) | 9.1 (1) |
| Black or African American | 5.7 (35) | 2.9 (3) | 5.6 (10) | 6.7 (15) | 7.5 (7) | 0 (0) |
| White | 78.7 (479) | 89.4 (93) | 78.1 (139) | 79.4 (177) | 66.7 (62) | 72.7 (8) |
| Asian or Asian American | 9.2 (56) | 8.7 (9) | 8.4 (15) | 9.4 (21) | 9.7 (9) | 18.2 (2) |
| American Indian or Alaska Native | 6.4 (39) | 2.9 (3) | 6.2 (11) | 6.3 (14) | 11.8 (11) | 0 (0) |
| Native Hawaiian or other Pacific Islander | 2.1 (13) | 2.9 (3) | 1.7 (3) | 1.8 (4) | 3.2 (3) | 0 (0) |

\* Multiple participants identified as 2 or more races.

### Assessment for ‘Bots’

The first bot detection question presented participants with a photo displaying a road number and name and prompted them to input the road name into a designated text box. Responses lacking either the road number or name were flagged as potential bots and subsequently excluded. The second bot detection approach employed questions about participants’ ages using two distinct formats: written and visual. Responses showing discrepancies exceeding a difference of 3 years or failing to match between formats were identified as potential bot entries and were excluded from analysis. A total of 864 complete, non-bot, survey responses were identified (Figure 1).

**Figure 1.**
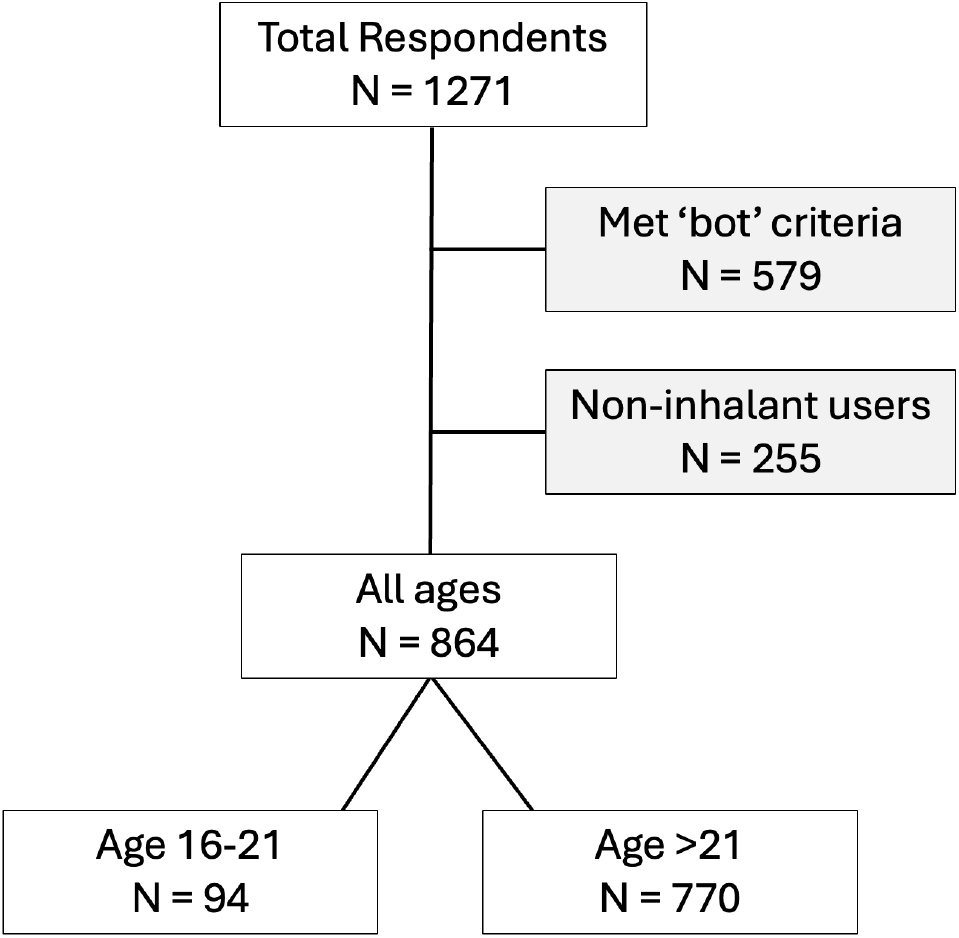
Breakdown of total survey responders who met criteria for inclusion into the study.

### Analysis

Raw data was exported into Microsoft Excel. Organized data was graphed and analyzed with GraphPad Prism (version 10.3.0; San Diego, CA).

## 3. Results

### 3.1. Rationale for Switching from Vaping Nicotine (using e-cigarettes) to Smoking Cigarettes

Out of 104 inhalant users who reported switching from vaping to smoking, 44% stated that they believed smoking was healthier than vaping (Figure 2a). Additionally, 40% noted that cigarettes were more accessible, and 38% mentioned having friends or family who smoke cigarettes as a factor in their decision. Among participants who provided only one reason for their switch, believing that smoking is healthier than vaping was the most common choice. None of the participants selected reasons such as social pressure, changing from sweet to harsher forms, normalization of smoking-related behaviors by e-cigarette vaping, or other miscellaneous reasons (Figure 2a).

**Figure 2.**
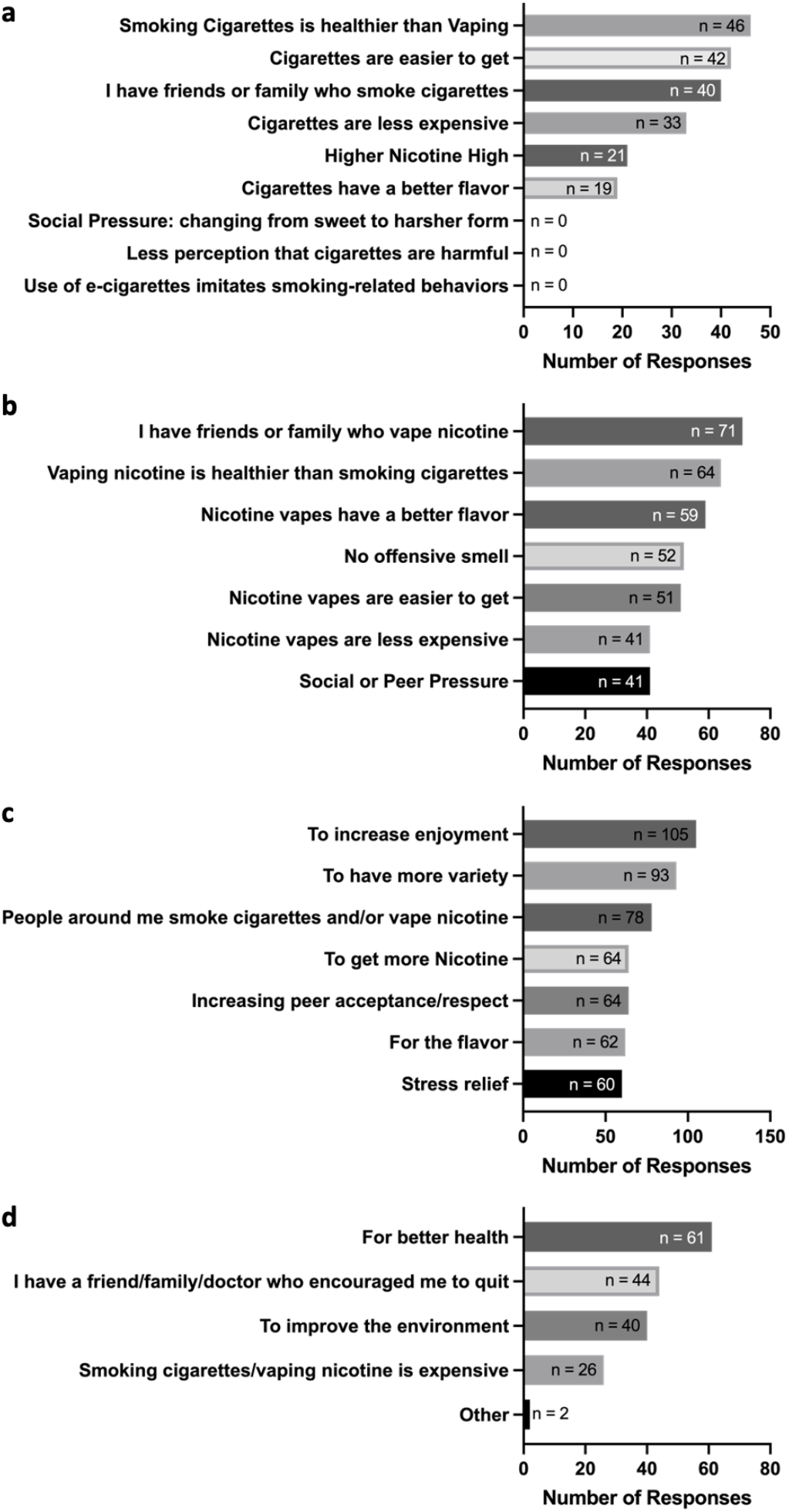
Drivers underlying switching from vaping to smoking, smoking to vaping, and multi-inhalant use. **a**. Main drivers of switching from vaping to smoking include the belief that smoking is healthier than vaping, the ease of obtaining cigarettes, and influence from friends or family who smoke. Notably, none of the respondents cited social pressure, the transition from sweet to harsh forms, reduced perception of harm, or the imitation of smoking-related behaviors by e-cigarettes as reasons for switching from vaping to smoking. **b**. The main drivers for transitioning from smoking to vaping were reported having friends or family who vape, the belief that vaping is healthier than smoking, and the perception that vapes have better flavor. **c**. The primary drivers for multi-inhalant use of vapes and combustibles include increasing enjoyment, having more variety, and being influenced by people around who smoke and/or vape. **d**. The main motivators behind quitting smoking and vaping were for better health, encouragement from friends, family, or doctors, and to improve the environment.

### 3.2 Rationale for Switching from Smoking Cigarettes to Vaping Nicotine

Among the 178 inhalant users who switched from smoking to vaping, 40% highlighted having friends or family who vape as a driver, while 36% expressed the belief that vaping is healthier than smoking (Figure 2b). Additionally, 33% cited the better flavor of vapes as a reason for their switch. Having a friend or family member who vapes was cited as a top reason among participants who selected only one reason for their switch (Figure 2b).

### 3.3 Multi-inhalant Use of Nicotine Vapes and Combustible Cigarettes

Among 223 participants who identified as multi-inhalant users, 47% (n = 105) highlighted increased enjoyment as the reason for multi-inhalant use, while 42% (n = 93) expressed a desire for more variety as a driver of this behavior (Figure 2c). Additionally, 35% (n = 78) reported having people around them who multi-inhalant use. Furthermore, 29% (n = 64) attributed their multi-inhalant usage to obtaining more nicotine, and 29% (n = 64) said they do it to increase peer acceptance or respect. Moreover, 28% (n = 62) mentioned that they smoke and vape because of the flavor, and 27% (n = 60) said they do it for stress relief. Participants who picked only one reason for multi-inhalant use chose either: 1. increasing enjoyment, 2. increasing peer acceptance or respect, and 3. flavor. No participants mentioned other reasons for their multi-inhalant use (Figure 2c).

### 3.4 Quitters of Tobacco Use

Out of the 93 participants who quit smoking and/or vaping, 66% (n = 61) attributed quitting to have better health, while 47% (n = 44) mentioned having a friend, family member, or doctor who encouraged them to quit as a contributing factor (Figure 2d). Additionally, 43% (n = 40) stated that it was to improve the environment, and 28% (n = 26) expressed that smoking and vaping are expensive. The top choice among participants who picked only one reason for quitting was better health. Notably, two participants provided other reasons – one mentioned that cigarettes are inferior, and the other did not specify (Figure 2d).

## 4. Discussion

In this study we explored reasons underlying transitions from vaping to smoking. Our findings revealed a nuanced picture, with multiple reasons contributing to switching from one inhalant to another (Infographic). Specifically, nicotine addiction did not emerge as the dominant factor. Instead, participants cited a variety of motivations, including beliefs that smoking is healthier than vaping, the influence of social networks, lower cost of cigarettes, and taste preferences. This multifaceted explanation challenges previous assumptions that nicotine addiction is the sole driver of transitioning from vaping to smoking.

An unexpected finding was the perception among some participants that smoking is healthier than vaping. This finding contradicts the prevailing literature, which broadly has determined that vaping is a less harmful alternative to smoking [9, 10]. This finding highlights a public health need for clear messaging about the known health effects of different forms of tobacco. While the public should be encouraged not to initiate e-cigarette vaping, as it does have adverse effects on health[6], those who already use combustible cigarettes should be shown the data demonstrating potential harm reduction of switching completely from smoking to vaping[7]. However, we are now finding that multi-inhalant use of both combustible tobacco and vaped nicotine has greater negative effects on health than either inhalant alone[8], and many smokers who add in e-cigarettes to reduce cigarette consumption end up as multi-inhalant users. Thus, the dangers of multi-inhalant use need to be clearly conveyed to the public.

In addressing our second research question as to why cigarette smokers switch to vaping nicotine, vaping being perceived as a healthier alternative, better flavors, and lack of offensive smell emerged as significant reasons. However, as with switching from vaping to smoking, our results revealed a diverse set of motivations for making the switch from smoking to vaping, including influence from social circles. Interestingly, reasons driving switching from smoking to vaping overlapped with those for transitioning from vaping to smoking, suggesting complex dynamics at play in individuals’ decisions regarding inhalant usage.

Multi-inhalant users of nicotine vapes and cigarettes were motivated by a desire for increased nicotine intake, as well as enjoyment, desire for variety, social influences, peer acceptance, flavor preferences, and stress relief. Both quitters of vaping and quitters of smoking citing concerns about their health, encouragement from friends, family, or healthcare providers, environmental considerations, and the cost of smoking/vaping as motivations for quitting. Additionally, eleven participants described their inhalant usage as occasional, particularly in social settings. While not fitting neatly into the categories of switching or quitting, these experiences offer insights into the context-specific nature of inhalant use and the role of social influences.

Limitations of the study include possible selection bias introduced through recruitment via social media. Additionally, the incentive offered for survey participation may have influenced participant motivation. Due to the need for brevity, longer surveys have lower click-through and completion rates[11], sociodemographic data beyond age, gender, race and ethnicity was not obtained. Future research should explore inhalant usage patterns in diverse populations.

## 5. Conclusions

There are complex reasons underlying inhalant use patterns, with a diverse array of motivations for switching, multi-inhalant use, and quitting smoking and vaping. These findings underscore the need for targeted interventions that address individual preferences, social influences, and perceptions of harm to effectively promote smoking and vaping cessation and harm reduction efforts. Further research is warranted to explore these dynamics in more depth and inform evidence-based strategies for addressing the public health challenges associated with smoking and vaping. Greater regulation of e-cigarette devices is needed to prevent initiation of vaping in adolescents and young adults, to prevent transitioning to combustible use or multi-inhalant use, both of which are known to cause more adverse health effects.

## Author Contributions

Conceptualization and methodology: EB, NK, KE, LCA. Formal analysis: EB, ZA, NK, HC, LCA. Investigation and data curation: EB, NK, HC, SP. Writing of the manuscript: all authors. Supervision: LCA; Funding acquisition: KE and LCA. All authors have read and agreed to the published version of the manuscript.

## Institutional Review Board Statement

The study was conducted in accordance with the Declaration of Helsinki, and approved by the Institutional Review Board of the University of California at San Diego (protocol 160204, approved 10/1/2023).

## Informed Consent Statement

Informed consent was obtained from all subjects involved in the study.

## Data Availability Statement

The raw data supporting the conclusions of this article will be made available by the authors on request.

